# Self-Management Competence in Depressed In-Patients: A Prospective Observational Study

**DOI:** 10.1101/2023.12.22.23300459

**Authors:** Nadja Schnierer, Iris Reinhard, Johanna Wagner-Dörr, Urs M. Nater, Matthias Bender, Peter M. Wehmeier

## Abstract

**Background:** Studies suggest that good self-management is associated with better coping with chronic mental conditions. However, an encompassing assessment of the relationship between depression and self-management competence is lacking.

**Methods:** This study assesses the relationship between depressiveness and self-management competence in a sample of 83 depressed in-patients. Beck Depression Inventory II (BDI-II) was used to assess depressiveness. The Self-Management Self-Test (SMST) was used to assess self-management competence. Patient surveys took place at the time of hospital admission (T1) and at the time of hospital discharge or approximately 4 to 6 weeks after (T2).

**Results:** Self-management competence correlated negatively with depressiveness at T1. Four out of five specific dimensions of self-management competence correlated inversely with depressiveness at T1. Self-management competence differed depending on the severity of the depressive syndrome and was higher the lower the severity of the depressive syndrome was. In the course of clinical treatment, self-management competence increased. Change of self-management competence during clinical treatment was not dependent on the sociodemographic variables gender or age. Change of self-management competence during clinical treatment predicted the change of depressiveness between hospital admission and discharge (T2 vs. T1) as well as depressiveness at T2.

**Limitations:** Sample size was relatively small. The SMST is a relatively new psychometric instrument that has not yet found widespread use in clinical research.

**Conclusion:** Our findings offer clinical evidence that in depressed in-patients, self-management competence and depressiveness are associated constructs. These results suggest that self-management competence may be a valuable resource in the treatment of depressive disorders.

## Introduction

Major depressive disorder (MDD) is a common illness, affecting more than 264 million people worldwide (James et al., 2018). The successful use of self-management strategies may reduce the symptom severity of depressive disorders (Levitt et al., 2009; Murphy et al., 2020). In relation to chronic medical conditions, self-management refers to the ability to cope with symptoms, treatments and physical and psychosocial effects related to the chronic condition (Corbin & Strauss, 1988; Barlow et al., 2002; Crepaz-Keay, 2010; Schulman-Green et al., 2012; Taylor et al., 2014). In the light of Antonovsky’s concept of resources (1979), depressive disorders can be considered as chronic conditions in which self-management competence is compromised (Barlow et al., 2002; Monroe & Harkness, 2012). Crepaz-Keay (2010) found that self-management strategies are still being underused in psychiatric patients despite a growing body of evidence for their effectiveness (Lean et al., 2019). However, a detailed and comprehensive investigation of the relationship between depression and self-management competence is lacking (Lean et al., 2019). To the best of our knowledge, this is one of the first studies ever conducted to explore this relationship.

## Materials and methods

### Instruments

The Self-Management Self-Test (SMST) was used to assess self-management competence (Wehmeier et al., 2019). The Beck Depression Inventory II (BDI-II) was used to assess depressiveness (Beck et al., 1996). The GAF scale (GAF) was used to assess the patientś level of psychosocial functioning (American Psychiatric Association, 1994).

### Procedure

*N* = 83 patients hospitalized for treatment of major depressive disorder in two psychiatric hospitals in a rural region in Central Germany filled out the SMST at the time of hospital admission (T1) and approximately 4 to 6 weeks later (T2), which in most cases equaled the day of hospital discharge. Additionally, patients were asked to fill out the BDI-II at T1 and T2. Experienced psychiatric clinicians filled out the GAF scale at T1. Data collection took place between April 2015 and September 2015.

### Ethics statement

The present study was approved in November 2014 by the Ethics Committee of the locally responsible Medical Board (Landesärztekammer Hessen FF 111/2014) and was carried out in full compliance with the Code of Ethics of the World Medical Association. Prior to their survey, all patients underwent an informed consent discussion during which they were informed about their right to interrupt their interrogation and withdraw their given consent at any time without having to give reasons therefor and without negative consequences. The patients were also informed that they could demand the deletion of their data at any time and without consequences. Additionally the patients were informed that their survey respectively their given answers would not be accessible to their psychiatrists and therapists in order to prevent their answers from affecting the patientś clinical treament. All patients received a written transcript of their rights and data protection laws and gave their written consent prior to participating in the study. Data was collected pseudonymized.

### Inclusion criteria

We included a clinical sample of *N* = 83 adults aged 18 to 65 years who had been hospitalized for the treatment of major depressive disorder according to ICD-10 GM criteria (WHO, 1993). Patients with severe mental disorders such as schizophrenia, manic episodes and bipolar disorder were excluded from the study, as mental disorders may impose an additional burden on one’s self-management competence and thus could be a source of bias. Secondly, patients with psychotic symptoms were excluded because psychotic symptoms may impair people’s self-management competence (Depp et al., 2016; Coventry et al., 2021), thereby possibly confounding the interpretation of treatment effects. For the same reasons, patients with severe physical illnesses were excluded.

### Statistical analyses

All statistical analyses were performed using SAS (SAS Institute Inc., 2013) or SPSS (IBM Corp., 2017, 2021). With regard to measures of central tendencies and variation, the statistical mean (*M*) ± its standard deviation (*SD*) is reported. The association of depressiveness and self-management competence in the recruited clinical sample was assessed by performing Pearson correlation analysis. Comparisons of the mean between two groups were tested by applying unpaired Student’s t tests. If more than two groups had to be compared, a one-way analysis of variance (ANOVA) including a Bonferroni-corrected post-hoc test was performed. For testing the difference between T2 and T1, a paired t test was applied. Linear regression analyses were used to predict the value of a dependent variable based on the value of one or more independent variable(s). With all statistical analyses, an alpha level of 0.05 was the basis for statistical significance.

### Aims of the study

This study assesses the association of depressiveness and self-management competence in depressed in-patients. Using the data from the clinical sample collected at T1 and T2, the following hypotheses were tested:

Hypothesis 1a: At T1, self-management competence correlated with the extent of depressiveness.
Hypothesis 1b: At T1, solitary dimensions of self-management correlated with depressiveness.
Hypothesis 2: At T1, the three levels of depression severity (Beck et al., 1996) differed in their average level of self-management competence from one another.
Hypothesis 3: The self-management competence of the patients increased during their clinical treatment.
Hypothesis 4: The change of self-management competence during clinical treatment was dependent on specific sociodemographic variables.
Hypothesis 5: The extent of change of self-management competence during clinical treatment (T2 vs. T1) predicted the change of depressiveness between T1 and T2.
Hypothesis 6: The extent of change of self-management competence during clinical treatment (T2 vs. T1) predicted depressiveness at T2.

## Results

### Patients’ characteristics

Eighty-two patients filled out the SMST at T1. Fourty-nine patients filled out the SMST at T2. One of these 49 patients filled out the SMST *only* at T2 (not at T1). Thus, the total sample size amounted to *N* = 83 patients.

Since only those patients were included in the statistical analyses for whom all required values were available (SMST and/or BDI-II), subsample sizes for each of the hypotheses tested differed from one another with regard to the number of patients included in the respective subsamples. A sample characterization based on sociodemographic variables can be found in **Table 1**.

**Table 1.** Patients’ characteristics at the time of hospital admission (T1).

Data on the patientś comorbidities could be obtained for 82 of the 83 patients of the total sample. Fifty percent of the total sample (i.e., 41 patients) suffered from one or more mental comorbidities. The most frequent mental comorbidities are listed in **Table 2**.

**Table 2.** Most frequent mental comorbidities in the total sample (*N* = 83).

Collectively, 48 SMST total scores at T1 *and* T2 were available (subsample *n*_3;4_). A graphical representation of the SMST total scores at T1 and T2 is displayed in **Figure 1**. Time-dependent BDI-II total scores (T2 vs. T1) are plotted in **Figure 2**.

**Figure 1.**
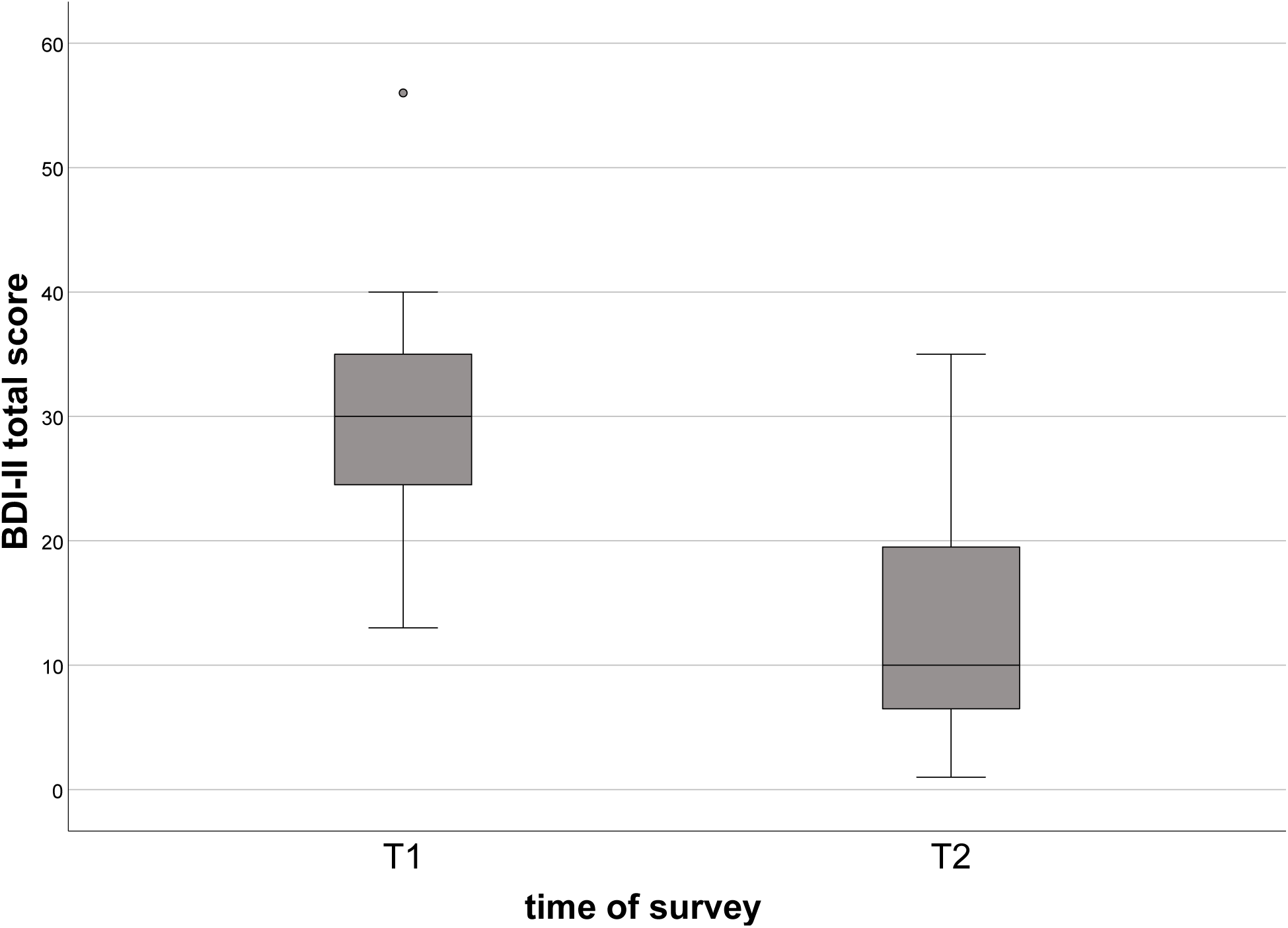
SMST total scores at T1 and at T2, respectively. *Note.* Subsample size *n*_3;4_ = 48. The boxes show the limits of the first and of the third quartile with the median in their centre (horizontal black line). The whiskers represent the range of the data with its maximum and its minimum. The empty dots above and below the left box (T1) are outliers.

**Figure 2.**
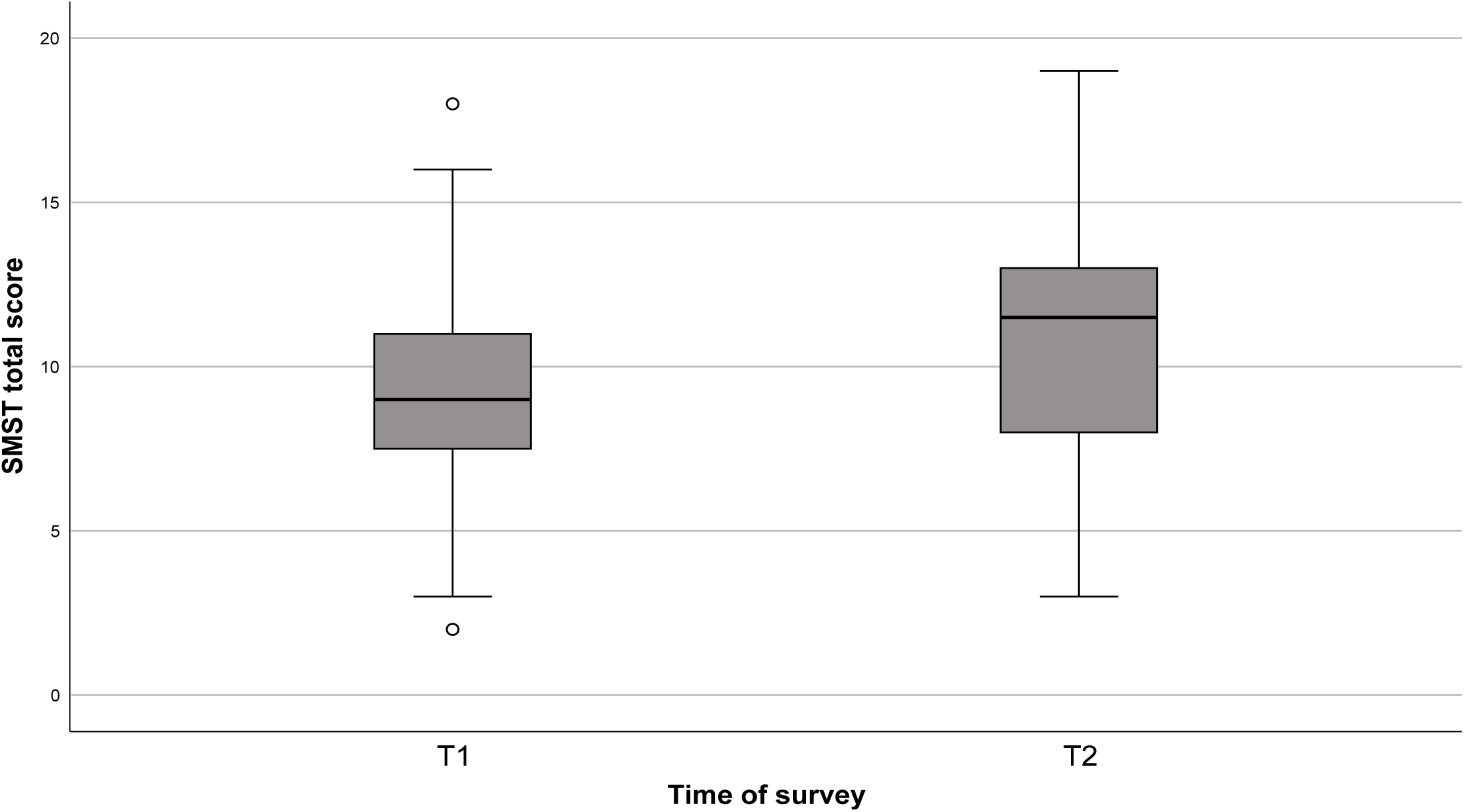
BDI-II total scores at T1 and at T2, respectively. *Note.* Subsample size *n3*;_4_ = 48. The gray boxes represent the limits of the first and of the third quartile with the median in their centre (horizontal black line). The whiskers show the scattering of the data. The gray dot above the left box (T1) is an outlier.

GAF scores (T1) were available for 58 of the 83 patients. At the time of hospital admission (T1), the mean GAF score of the sample was *M* = 36.8 [%], *SD* = 10.1 [%].

### Hypothesis 1a. Correlation of self-management competence and the level of depressiveness at the time of hospital admission (T1)

From a total of *n*_1_ = 52 patients, both a SMST total score and a BDI-II total score were available at T1. Beck Depression Inventory II (BDI-II) was used to assess depressiveness. The Self-Management Self-Test (SMST) was used to assess self-management competence. The association between these two variables was assessed performing Pearson correlation analysis. With the correlation coefficient being *r* = -.50 (*p* < .001), the BDI-II total score and the SMST total score showed a statistically significant, strong correlation at T1 (Cohen, 1988). Accordingly, a high level of self-management competence at the time of hospital admission (T1) was associated with a low level of depressivenes and vice versa. Thus, the hypothesis that the SMST total score (which reflects the individual self-management competence) and the BDI-II total score (which is a measure of depressiveness) correlated with one another at the time of hospital admission could be confirmed.

### Hypothesis 1b. Correlation of solitary dimensions of self-management with depressiveness at T1

As mentioned above, subsample size for the statistical analysis was *n*_1_ = 52. The five items of the SMST reflect the five dimensions of self-management according to Wehmeier (2015). As with the calculations for Hypothesis 1a, the linear association between the two variables was assessed performing Pearson correlation analysis. **Table 3** below shows the corresponding correlation matrix. We found that a high degree of depressiveness at the time of hospital admission (T1) was associated with

a. low value for the self-management dimension *awareness*,
b. a low value for the self-management dimension *planning*,
c. a low value for the self-management dimension *decision-making*,
d. a low value for the self-management dimension *action* and vice versa.

**Table 3.** Means, standard deviations and correlations between solitary SMST items, SMST total score and BDI-II total score at the time of hospital admission.

The self-management dimension *relationship*s did not show a significant correlation with depressiveness at T1 (p > .05).

Thus, the hypothesis that solitary dimensions of self-management correlated with depressiveness at the time of hospital admission was confirmed for four of the five dimensions of self-management according to Wehmeier et al. (2019).

### Hypothesis 2. Differences in the average level of self-management competence in dependence on the level of depression severity at T1

We conducted a one-factorial analysis of variance (ANOVA) in order to assess whether there was a statistically significant difference in the average level of self-management competence dependent on the level of depression severity at T1. Depression severity levels were categorized according to the BDI-II total score as specified by Beck et al. (1996).

From a total of *n*_1_ = 52 patients, both a SMST total score and a BDI-II total score were available at T1 (see hypotheses 1a and 1b). Two of these 52 patients presented with a BDI-II total score of ≤ 10 points at T1 and could therefore not be assigned to the three degrees of depression severity *mild*, *moderate* or *severe* according to Beck et al. (1996). After excluding the two patients mentioned above, subsample size for the ANOVA was *n*_2_ = 50.

Analysis revealed a statistically significant mean difference between the three groups examined, *F*(2, 47) = 3.90, *p* = .027. However, a Bonferroni-corrected post-hoc-test showed a statistically significant difference only for the two groups *mild depressive syndrome* (*M* = 11.3, *SD* = 4.0) and *severe depressive syndrome* (*M* = 8.0, *SD* = 2.9). The group of patients suffering from a *moderate depressive syndrome* (*M* = 9.5, *SD* = 2.7) was not significantly different from the two groups mentioned before after the Bonferroni-corrected post-hoc-test.

Thus, the hypothesis that at T1, the three levels of depression severity differed in their average level of self-management competence from one another was confirmed only for the groups suffering from a *mild* depressive syndrome and a *severe* depressive syndrome. The hypothesis could not be confirmed for the group of patients suffering from a *moderate* depressive syndrome.

### Hypothesis 3. Increase of the self-management competence of the patients during their clinical treatment

Collectively, 48 SMST total scores at T1 *and* T2 were available (*n*_3;4_). We calculated the difference Δ SMST (T2-T1) as a variable measuring the change in self-management competence during clinical treatment. Figure 1 shows the SMST total scores at T1 and T2. At T1, the SMST total score was *M*_1_ = 9.0, *SD*_1_ = 0.5. At T2, the SMST total score was *M*_2_ = 10.9, *SD*_2_ = 0.5. A paired t-test showed that the increase in self-management competence in the course of the clinical treatment was statistically significant, *t*(47) = ™3.00, *p* = .004.

### Hypothesis 4. Dependency of the change of self-management competence during clinical treatment on sociodemographic variables

#### Gender

An unpaired t-test was used to examine whether men and women had improved to different extents in their self-management competence during clinical treatment. The variable *gender* was the independent variable in the analysis. The variable *change in self-management competence during clinical treatment* was the dependent variable. The descriptives are reported in **Table 4**. The variable *change in self-management competence during clinical treatment* (equalling Δ SMST) showed no statistically significant gender-related difference, *t* (46) = - .76, *p* = .454, *n*_3;4_ = 48.

**Table 4.** Gender difference for Δ SMST (T2-T1) in subsample *n*_3;4_.

#### Age

Pearson correlation analysis showed no statistically significant correlation between the variables *age* and *change in self-management competence during clinical treatment*, *r* = .03, *p* = .862, *n*_3;4_ = 48. Thus, the hypothesis that the change in self-management competence in the course of clinical treatment was dependent on specific sociodemographic variables was not confirmed for the variables *gender* or *age*.

### Hypothesis 5. Prediction of the change of depressiveness during clinical treatment by the extent of change in self-management competence during clinical treatment

The statistical relationship between the predictor (*x*) *change in self-management competence during clinical treatment* (Δ SMST (T2-T1)) and the criterion (*y*) *change in depressiveness during clinical treatment* (Δ BDI (T2-T1)) was assessed using simple linear regression analysis. From a total of *n*_5;6_ = 23 patients, BDI-II total scores and SMST total scores were available at T1 and T2. **Table 5** reports the results of the analysis. In the subsample examined (*n*_5;6_), the change in self-management competence during the clinical treatment was a significant predictor for the change in depressiveness during clinical treatment (*b* = ™1.10, *p* = .022). Thus, the hypothesis that an increase in self-management competence would correlate with a decrease in depressiveness during clinical treatment was confirmed.

**Table 5.** Regression analysis results using *change in depressiveness between hospital admission and discharge (T2 vs. T1)* as the dependent variable.

### Hypothesis 6. Prediction of depressiveness at T2 by the extent of change in self-management competence during clinical treatment

Multiple linear regression analysis was performed in order to assess the association between the two predictors (*x*_i_) *change in self-management competence between T1 and T2* (Δ SMST (T2-T1)) and *depressiveness at T1* with the criterion (*y*) *depressiveness at T2*. For subsample size and calculation of the difference Δ SMST see *hypothesis 5*. **Table 6** reports the results of the analysis. In summary, the change in self-management competence during clinical treatment was a statistically significant, negative predictor for depressiveness at T2 in the subsample examined (*b* = ™1.06, *p* = .008, *n*_5;6_ = 23). The higher the change was in SMST, the lower was the BDI-II at T2.

**Table 6.** Regression analysis results using *depressiveness at T2 (hospital discharge)* as the dependent variable.

Depressiveness at T1 was a statistically significant, positive predictor for depressiveness at T2 (*b* = .40, *p* = .028, *n*_5;6_ = 23). The higher the BDI-II was at T1, the higher was depressiveness at T2. The change in self-management competence played a slightly greater role for predicting depressiveness at T2 than depressiveness at T1. When controlling for the additional variables *gender* and *age*, the two predictors (*x*_i_) mentioned above remained statistically significant, whereas the additional variables *gender* and *age* showed no statistically significant influence. Thus, the hypothesis that the extent of change in self-management competence during clinical treatment would predict depressiveness at the time of the second survey was confirmed.

## Discussion

It is widely known that depressed patients may suffer from functional impairments in various areas of everyday life and in personal abilities and skills (McKnight & Kashdan, 2009). Skills that are known to be affected by depressive disorders and to be impaired in depressed patients include cognitive or executive skills, e.g. (Murphy et al., 2001; Fossati et al., 2002; Royall et al., 2002; DeBattista, 2005; Yen et al al., 2011). Cognitive impairments, inhibition of drive, fatigue, exhaustion and feelings of overload can negatively influence the patientś cognitive abilities, planning capabilities, decision-making abilities, as well as the abilities to act and interpersonal abilities (Iosifescu, 2012; DGPPN et al., 2017). Thus, we expected the extent of depressive disorder of the clinical sample to have a negative impact on the five dimensions of self-management according to Wehmeier et al. (2019), being *awareness*, *relationships*, *planning*, *decision-making* and *action*. Furthermore, we expected that impairments like the ones described above would be more severe with higher levels of depressiveness.

Our results confirm this assumption (**hypothesis 1a**) and indicate a correlation between the two variables examined, i.e. the patients’ individual self-management competence and their level of depressiveness at the time of hospital admission (T1). Additionally, statistical analyses confirmed that the severity of the depressive disorder of the patients were negatively correlated with four of the five dimensions of self-management according to Wehmeier et al. (2019), i.e. awareness, planning, decision-making and action (**hypothesis 1b**). In accordance with these results, Leykin et al. (2011) found that the ability to make decisions is being impaired by depressive disorders.

Thus, our study confirms findings from previous studies in patients with depression (Weingartner et al., 1981; Iosifescu, 2012; Rock et al., 2014). Nevertheless, there was no statistically significant correlation between the SMST item *relationships* and depressiveness at the time of hospital admission. This result does not confirm the observations from previous depression research (Coyne, 1976; Murphy, 1985; Joiner, 1999; Ge et al., 2017). This finding may have resulted from the limited sample size.

However, the sample size of *N* = 83 was large enough to answer the research questions of the study. The fact that the SMST item *relationships* did not reach statistical significance may have been due to the large proportion of patients whose marital status was *single*, *divorced/living apart* or *widowed* (**Table 1**). This subgroup of patients may have had no problems arising in their interpersonal relationships shortly before or at the time of hospital admission *or* they may have played a smaller role than the other four self-management dimensions. Furthermore, the fact that depressive disorders are often accompanied by social withdrawal or social isolation could have culminated in a skewed self-assessment of the patientś interpersonal competence (aggravation or dissimulation of competence) or in a recall bias when retrospectively assessing interpersonal competence.

We also found that the extent of depressiveness or rather the severity of the depressive syndrome seems to be associated with the patientś self-management competence (**hypothesis 2**). As shown by Orem (2001), Fox (2017), Krijnen-de Bruin (2021) and in our previous work Wehmeier et al. (2019), the ability of patients to make use of their self-management skills is influenced by the current severity of their depressive symptoms, with a higher symptom severity leading to a decrease in self-management skills. The present study allows a similar conclusion. The average self-management competence of the clinical sample differed between groups of patients according to depression severity with a *severe* depressive syndrome leading to a lower level of self-management competence than a *mild* depressive syndrome. In the present study the self-management competence of patients suffering from a *moderate* depressive syndrome did not differ from those suffering from either a *mild* depressive syndrome or a *severe* depressive syndrome (**hypothesis 2**).

In the present study, the patients’ self-management competence increased in the course of clinical treatment (**hypothesis 3**). This result met our previous expectations. Given the fact that in functional neuroimaging, psychotherapeutic interventions have been shown to have regulatory effects on dysregulated cerebral activity patterns in depressed patients (Beck et al., 1979; DeRubeis et al., 2008; Kuhn et al., 2014; Straub et al., 2015; Han et al., 2018), we assumed that an increase in the patientś self-management competence would be accompanied by a modification of dysregulated activity in affected cerebral circuits. Not only because of the well-known positive effect that psychopharmacological and psychotherapeutic interventions exert on depressive disorders (Schneider et al., 2005; Liebherz & Rabung, 2013; Bundesärztekammer et al., 2022), the increase in self-management competence observed in our study can be considered a direct consequence of clinical treatment.

Fourthly, the increase in self-management competence was found not to depend on age or gender (**hypothesis 4**). Previous research in physical disorders has demonstrated age-related and gender-related differences in self-management competence (Ruggiero et al., 1997; Brewer-Lowry et al., 2010; Burner et al., 2013; Shrestha et al., 2013; Hornung-Praehauser et al., 2016). However, in mental disorders evidence is limited (Lean et al., 2019). An assessment of the effectiveness of cooperative models of care for chronic mental disorders which comprise *patient self-management support* concludes that there are neither age-related nor gender-related differences in clinical effectiveness of these programs (Miller et al., 2013). Our findings reflect these results. Future research should investigate this topic further and focus on examining a larger sample in order to provide a solid evidence base for making informed decision on future treatment strategies.

In our sample, the change in self-management competence in the course of the clinical treatment was a significant predictor for the change in depressiveness in the same period of time (**hypothesis 5**). The extent of change in self-management competence predicted depressiveness at T2, i.e. at the time of hospital discharge or shortly thereafter (**hypothesis 6**).

The extent of change in self-management competence played a slightly greater role in predicting depressiveness at T2 than the extent of depressiveness at hospital admission (**hypothesis 6**). This finding is not only of value for psychotherapists involved in the treatment of depressed patients but also for the patients themselves whose expectation of self-efficacy can be strengthened when being presented with this result of our study.

Previous research has shown (a) that self-management programs and the use of self-management strategies both may positively affect depressed patients and (b) that the severity of depressive symptoms can be mitigated by the successful use of self-management strategies (Ludman et al., 2003; Levitt et al., 2009; Ryan et al., 2010; Villaggi et al., 2015; van Grieken et al., 2018; Murphy et al., 2020). In the therapy of chronic somatic diseases, interventions enhancing self-management competence have been shown to be effective in reducing comorbid depressive symptoms (Kotses et al., 1995; Barlow et al., 2000; Lorig et al., 2001; Moon et al., 2018; Whitebird et al., 2018).

Selfweersonal resource which can be developed, shaped and actively worked on during psychotherapeutic interventions (Murphy et al., 2020). Our finding that an increase in self-management competence led to a decrease in depressiveness in our clinical sample (**hypothesis 5**) implies that the (psychotherapeutic) interventions to improve self-management competence of depressed patients hold significant potential to further promote the recovery of patients with depressive disorders. In line with the work of Murphy et al. (2020), our results suggest that self-management competence can be a powerful resource for recovery from mental disorders. Thus, self-management competence as a resource could be an important and effective component in the treatment of depressive disorders.

### Limitations

Limitations of our study include the limited sample size and the limited geographical area served by the hospitals we recruited the study participants at. We used the self-report scale BDI-II to assess the depressiveness of the patients. Clinical assessment of the patients according to ICD-10 criteria was provided by the clinician. The second limitation of our study arises from the fact that no additional diagnostic instruments had been used to validate the clinical diagnosis. Future research on this topic should confirm clinical diagnoses with a diagnostic inventory for mental disorders, such as the SCID-5- CV or the SCID-5-PD. Thirdly, the specific clinical interventions received by the patients during their clinical treatment (e. g., pharmacological, psychotherapeutic) were not recorded because the specific type and extent of clinical interventions were not part of the study design. They were not relevant for testing the hypotheses that were investigated. However, all patients received treatment according to current guidelines as well as international standards of care. Lastly, we assessed self-management competence by using the Self-Management Self-Test (SMST), which is a relatively new psychometric instrument that has not yet found widespread use in clinical research. However, the SMST is currently being used in a large online psychotherapy study in patients with depression (Baumeister et al., 2021).

## Conclusion

The present study investigates the association of depressiveness and self-management competence in a clinical sample of depressed in-patients. To date, individual self-management competence has played a minor role in clinical research and in the therapy of depressive disorders. Our findings offer clinical evidence that in depressed in-patients, self-management competence and depressiveness are associated constructs. These results suggest that self-management competence is a valuable resource well worth enhancing in the treatment of depressive disorders.

## Data Availability

All relevant data are within the manuscript and its Supporting Information files.

## Acknowledgements

We thank all participants of this study and all research collaborators for their invaluable work and advice in data analysis and their support in the process of creating this work.

